# Lessons drawn from China and South Korea for managing COVID-19 epidemic: insights from a comparative modeling study

**DOI:** 10.1101/2020.03.09.20033464

**Authors:** Biao Tang, Fan Xia, Nicola Luigi Bragazzi, Xia Wang, Sha He, Xiaodan Sun, Sanyi Tang, Yanni Xiao, Jianhong Wu

**Affiliations:** The Interdisplinary Research Center for Mathematics and Life Sciences, Xi’an Jiaotong University, Xi’an 710049, People’s Republic of China; Laboratory for Industrial and Applied Mathematics, Department of Mathematics and Statistics, York University, Toronto, Ontario, Canada, M3J 1P3; School of Mathematics and Information Science, Shaanxi Normal University, Xi’an, 710119, People’s Republic of China; School of Mathematics and Statistics, Xi’an Jiaotong University, Xi’an 710049, People’s Republic of China; Fields-CQAM Laboratory of Mathematics for Public Health, York University, Toronto, Ontario, Canada, M3J 1P3

**Keywords:** COVID-19 epidemic, multi-source data, mathematical model, mainland China and South Korea, comparative study

## Abstract

We conducted a comparative study of COVID-19 epidemic in three different settings: mainland China, the Guangdong province of China and South Korea, by formulating two disease transmission dynamics models incorporating epidemic characteristics and setting-specific interventions, and fitting the models to multi-source data to identify initial and effective reproduction numbers and evaluate effectiveness of interventions. We estimated the initial basic reproduction number for South Korea, the Guangdong province and mainland China as 2.6 (95% confidence interval (CI): (2.5, 2.7)), 3.0 (95%CI: (2.6, 3.3)) and 3.8 (95%CI: (3.5,4.2)), respectively, given a serial interval with mean of 5 days with standard deviation of 3 days. We found that the effective reproduction number for the Guangdong province and mainland China has fallen below the threshold 1 since February 8^th^ and 18^th^ respectively, while the effective reproduction number for South Korea remains high, suggesting that the interventions implemented need to be enhanced in order to halt further infections. We also project the epidemic trend in South Korea under different scenarios where a portion or the entirety of the integrated package of interventions in China is used. We show that a coherent and integrated approach with stringent public health interventions is the key to the success of containing the epidemic in China and specially its provinces outside its epicenter, and we show that this approach can also be effective to mitigate the burden of the COVID-19 epidemic in South Korea. The experience of outbreak control in mainland China should be a guiding reference for the rest of the world including South Korea.

## Introduction

Coronavirus, an enveloped virus characterized by a single-stranded, positive-sense RNA, causes generally mild infections but occasionally lethal communicable disorders leading to SARS, MERS^1^ and the current COVID-19 outbreak^2,3^ that has gradually spread out from the epicenter Wuhan/China and affected 103 countries/territories and international conveyances including the cruise ship Diamond Princess harbored in Yokohama/Japan as of March 7^th^ 2020.

In the absence of effective treatments and vaccines, an early adoption of stringent public health measures is crucial in mitigating the scale and burden of an outbreak. Unprecedented restrictive measures, including travel restrictions, contact tracing, quarantine and lock-down of entire towns/cities adopted by the Chinese authorities have resulted in a significant reduction of the effective reproductive number of COVID-19^4,5^. However, these public health interventions may not be considered and/or implemented as effectively in other settings and contexts. Decision-making and implementations may require adaptations and modifications to take into account setting-specific characteristics in terms of community features, local epidemiology and risk assessment, social habits, juridical provisions, organizational coordination, and availability of economic-financial resources. For instance, particularly restrictive measures may not be effective in certain countries^6^.

Several public health interventions can be implemented to counteract the threat posed by an emerging outbreak^7^ with pandemic potential. These interventions can be basically classified into two major categories: the measures of the first category are aimed at protecting the borders and include interventions like travel restrictions and border entry screening, whereas the measures of the second category have the objective of locally controlling the spreading of the virus and include enhanced epidemiological surveys and surveillance, contact tracing, school closure and other interventions that favor a reduction in number of social contacts. The effectiveness of such measures from both group is variable and some is still under debate. Regarding, for example, extensive travel restrictions, a recent systematic review has shown that this intervention may contribute to delaying but not preventing the transmission and diffusion of a viral outbreak. As such, it is not recommended for implementation, if not within a broader package of public health measures aimed at rapidly containing the outbreak^8^. A similar conclusion can be reached for border entry screening, considered as ineffective or poorly effective *per se*, and therefore needs to be combined and provided together with other strategies^9^. School closure appears to be potentially effective in containing/reducing viral outbreaks, although further research is warranted to identify the best strategy in terms of timing and length of closure^10^. The measure of quarantine is also particularly controversial, since it raises ethical dilemmas, and political and social concerns^11,12^ and quantification of its real impact^11^ is difficult due to a high uncertainty in its efficacy. However, in the absence of effective medical interventions, these measures must be implemented and the success of these measures, despite their disruptive impact on social-economic activities, depends heavily on how these measures are adapted to the specific scenario, in terms not only of clinical and epidemiological variables but also of social aspects, including social habits, juridical provisions, and economic-financial resources.

How differentiation and combination of these interventions within a coherent and systematic package of public health measures contributes to different outbreak outcomes is an urgent global health issue that must be addressed in order to ensure that lessons from countries that have early experienced COVID-19 outbreak can be learnt by other countries in their preparedness and management of a likely pandemic.

In South Korea, the first COVID-19 case (a 36 years old Chinese woman, with a recent travel history to Wuhan) was reported on January 8^th^ 2020. A severe cluster of cases emerged in the city of Daegu, where on February 23^rd^ 2020 a 61 years old woman spread the virus to hundreds of worshippers at Shincheonji Church of Jesus. On March 5^th^ 2020, a further cluster of cases occurred at a nursing home in Gyeongsan, which has been declared “special care zone” in an effort to contain the viral outbreak. As of March 8^th^ 2020, South Korea has reported 7,313 cases, with 130 total recovered cases and 50 deaths, with no sign that the epidemic is slowing down.

In comparison, intensive social contacts and massive mobility associated with the Chinese Traditional Spring Festival, combined with an initial delay in responding to the outbreak, resulted in an exponential growth of infections in the (then) epicenter (Wuhan) and large case importations to other Chinese cities. On January 23^rd^ 2020, the Chinese government decisively implemented a systematic package of measures in the epicenter, including the lock-down/quarantine of Wuhan city and other cities/towns of the Hubei province, intense contact tracing and isolation. This led to rapid and effective mitigation of the COVID-19 epidemic. Case importation before the January 23^rd^ lock-down also resulted in outbreaks in all Chinese provinces, but the systematic package of interventions implemented across the country led to effective containment. On March 8^th^ 2020, the newly confirmed cases in the entire country reduced to 40.

Particularly, Guangdong, the province with the largest population in China at present, with GDP ranked first since 1989 and with the level of middle and upper income countries and middle developed countries, reported the first confirmed COVID-19 case on January 19^th^. On January 23^rd^, the government of Guangdong province announced the first-level response to major public health emergencies for controlling the spread of COVID-19. As of March 8^th^, there are totally 1325 confirmed cases and 8 deaths in Guangdong province, and no new case is reported. In contrast to South Korea, there was a relatively large ratio of imported cases in Guangdong province, particularly, in Shenzhen (a mega city in Guangdong province) more than 70 percent of the confirmed cases are imported^13^.

What differences in the intervention design and implementation between China and South Korea, and between Korea and Guangdong have contributed to the different epidemic curves, and what projection can we make about the epidemic trend in South Korea under different scenarios if a portion or the entirety of the Chinese package of public health interventions is applied to the South Korea? These are the questions we aim to answer by developing mathematical models tailored to different settings, including the entire country of China and its Guangdong province and South Korea.

## Methods

### Data

We obtained data of the confirmed COVID-19 cases, cumulative number of quarantined individuals, cumulative death cases in mainland China from the “National Health Commission” of the People’s Republic of China^14^. Data information includes the newly reported cases, the cumulative number of reported confirmed cases, the cumulative number of cured cases and the number of death cases, as shown in Figure 1(A-C). In addition, we obtained the data of the cumulative confirmed cases, cumulative cured cases and daily cases under medical observation for the Guangdong province (Figure 1(E)) of China. We also obtained the data of cumulative confirmed cases and cumulative tested cases for South Korea from the Korea Centers for Disease Control and Prevention (KCDC)^15,16^, as shown in Figure 1(D), (F). The data were released and analyzed anonymously. Note that the first confirmed case was reported on January 23^rd^ 2020 for South Korea, and also on January 23^rd^ 2020 mainland China started the lock-down of Wuhan city, the epicenter, and implemented other interventions. Note that the data for reported cases, either confirmed or quarantined, or under medical observation or tested, was used in China or South Korea since January 23^rd^ 2020.

**Figure 1.**
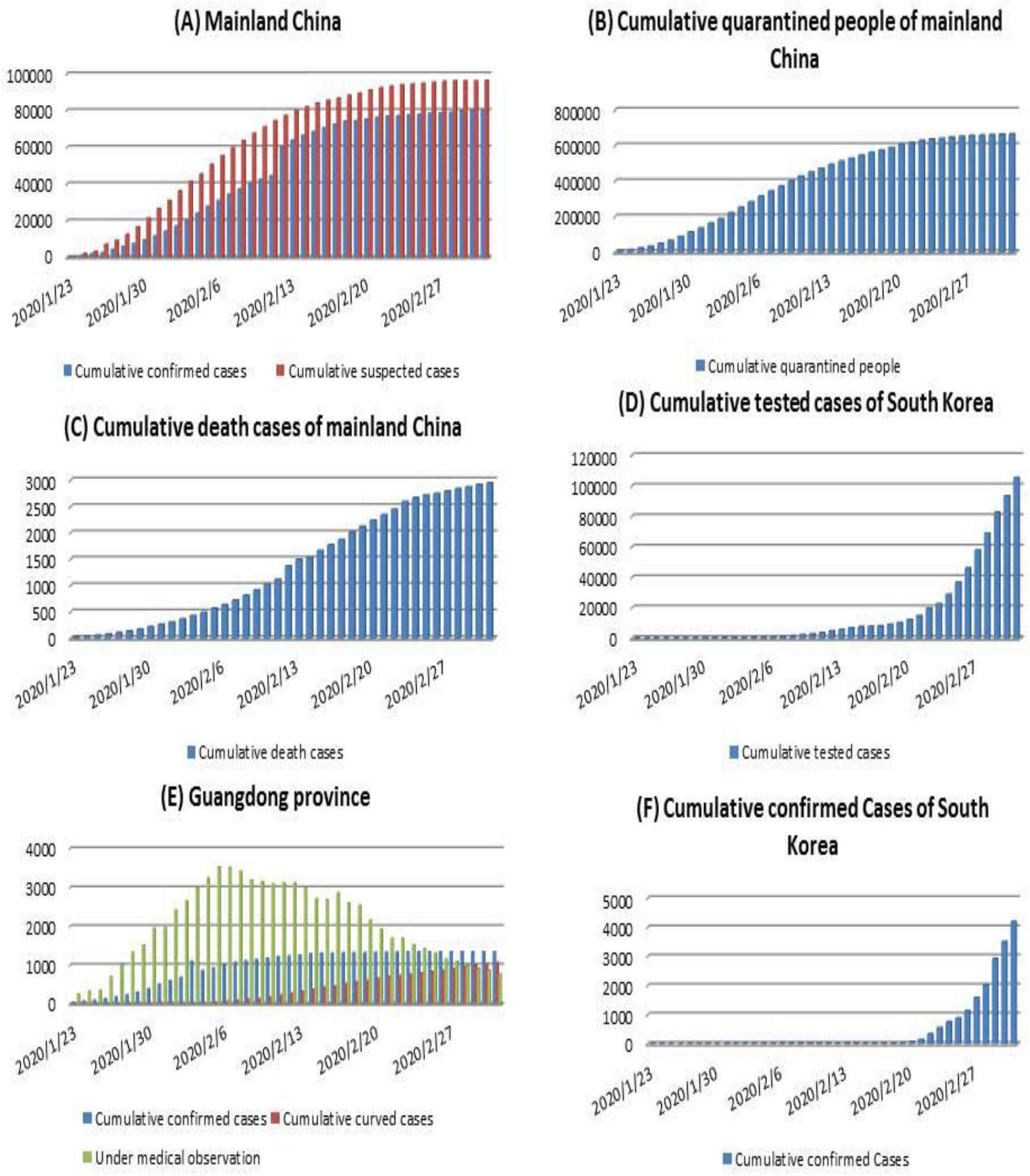
The datasets related to the COVID-19 epidemics, including newly reported cases, cumulative number of reported cases, cumulative number of cured cases, cumulative number of death cases, cumulative quarantined cases and cumulative suspected cases for mainland China (A-C), the Guangdong province of China (E), and South Korea (F).

### The model

Our baseline model is the classical deterministic susceptible-exposed-infectious-removed (SEIR) epidemic model refined by incorporating contact tracing-quarantine-test-isolation strategies (Figure 2). We stratify the population into susceptible (*S*), exposed (*E*), symptomatic/asymptomatic infected (*I*/ *A*), hospitalized (*H*) and recovered (*R*) compartments, and we further stratify the population to include quarantined susceptible (*S*_*q*_), and quarantined suspected individuals (*T*). These stratifications were used in our previous studies^4,5,17^ and agreement of model predictions with real data provides a validation of the model structure reflecting the interventions implemented in Wuhan and in mainland China. Here, we add an additional quarantined suspected compartment, which consists of exposed infectious individuals resulting from contact tracing and individuals with common fever. These individuals with common fever but quarantined as COVID-19 suspected contributed to the difficulty of implementing an effective quarantine process due to the size of this compartment. In what follows, exposure, transmission and infection compartments are always used for modeling the COVID-19.

**Figure 2:**
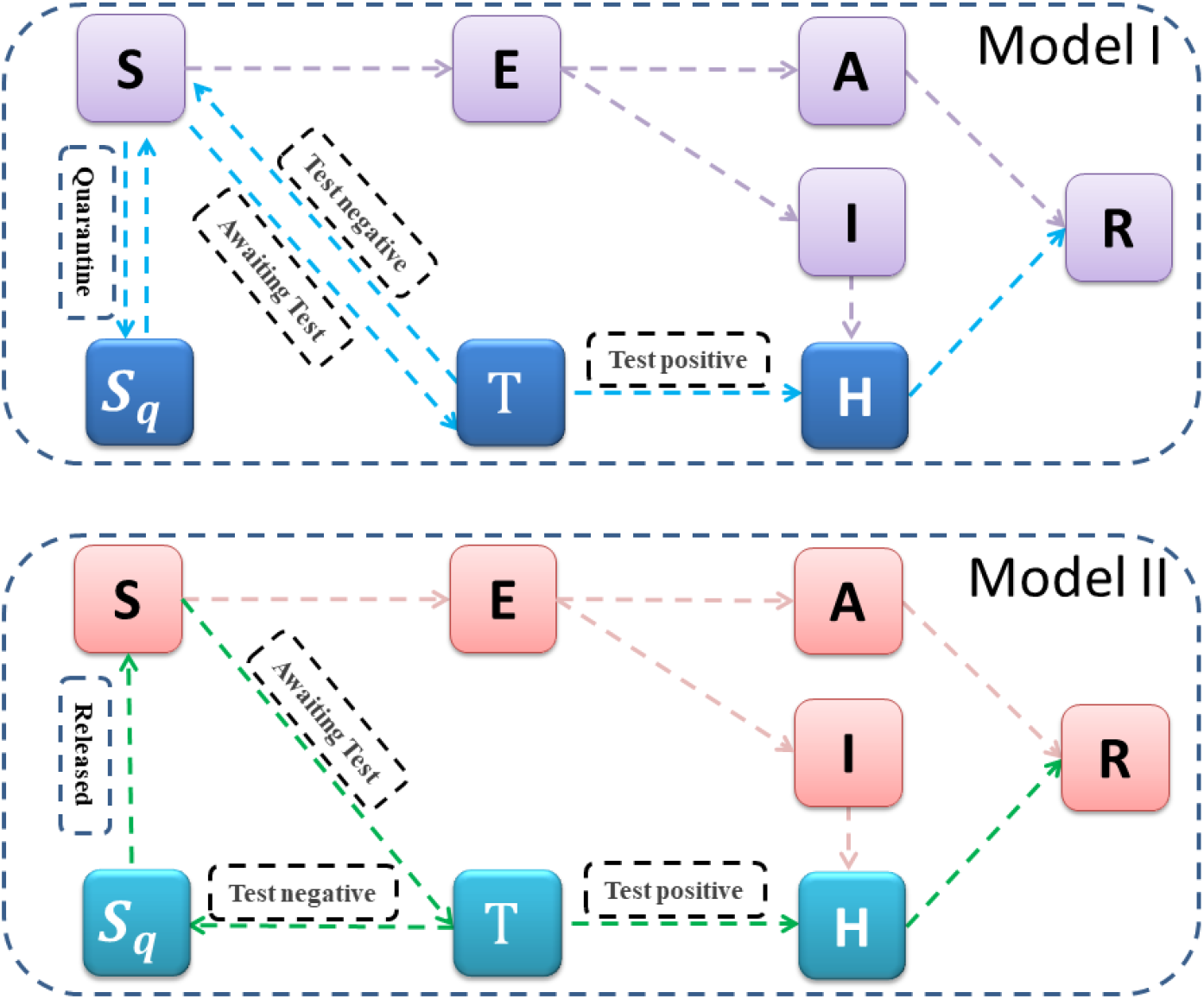
The illustration of the compartmental models incorporating important interventions and features of reporting systems, for mainland China (model I) including its Province of Guangdong, and for South Korea (model II).

In our model formulation, the transmission probability is denoted by β and the contact rate is denoted by c. By enforcing contact tracing, a proportion, *q*, of individuals exposed is quarantined, and can either move to the compartment *B* or *S*_*q*_ with rate of β*cq* (or (1 – β)*cq*)), depending on whether they are effectively infected or not^18,19^, while the other proportion, *1 – q*, consists of individuals exposed to the virus who are missed from contact tracing and move to the exposed compartment *E* at rate of β*c*(1 - *q*) once effectively infected or stay in compartment *S* otherwise. Note that contact tracing is not triggered by asymptomatic infected individuals, who can infect susceptible individuals. We use constant *m* to denote the transition rate from the susceptible compartment to the (COVID-19) suspected compartment due to fever and/or illness-like symptoms. The suspected individuals leave this compartment at a rate *b*, with a proportion, *f*, if confirmed to be infected with COVID-19, going to the hospitalized compartment, whilst the other proportion, 1-*f*, is ruled out for COVID-19 infection and goes back to the susceptible compartment. Then, for mainland China, the model equations are as follows.

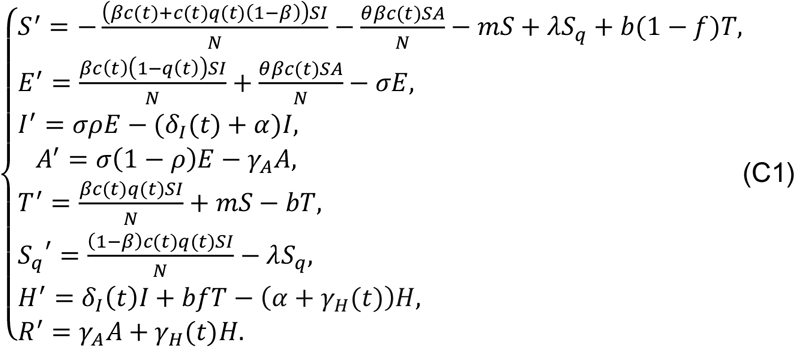

The effective reproduction number is given by

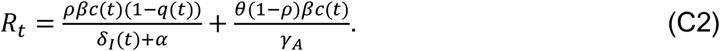

In epidemiology, the basic reproduction number (R_0_), used to measure the transmission potential of a disease, is the average number of secondary infections produced by a typical case of an infection in a population where everyone is susceptible. Once the intervention implemented results effective, the effective transmission potential is measured by the effective reproductive number *R*_*t*_ that can be time-dependent as intervention measures vary over time. If *R*_*t*_ falls below 1 and remains below 1, there will be a decline in the number of cases.

The prevention and control interventions were gradually improved in mainland China, and there are several key time points when mitigation measures were gradually strengthened: 1) On January 23^rd^ Wuhan was locked down, and most parts of China shortly adopted a similar strategy; 2) On January 26^th^, the government announced to extend the Chinese Traditional New Year Festival holiday so self-isolation/protection was maximized; 3) On February 7^th^ the Chinese government created the partnership between each one of the 16 provinces to its sister-city in the epicenter, the Hubei province, to reinforce the health care workers and equipment in the sister-city in Hubei; 4) On February 12^th^ the Hubei province started to include the clinically diagnosed cases into the confirmed cases to enhance its quarantine/isolation measure; 5) On February 14^th^, Wuhan refined its management protocol of residential quarters; 6) On February 16^th^, the National Health Commission of the People’s Republic of China revised its New Coronavirus Pneumonia Prevention and Control Plan to further clarify and enhance the public health interventions in four key areas: Quarantine high-risk individuals as much as possible, Test suspected individuals as much as possible; Treat patients as best as possible; and Receive and cure all the patients. We describe this improvement by using the time-dependent contact rate *c*(*t*), quarantined rate *q*(*t*), and detection rate *δ*_*I*_ (*t*), as follows

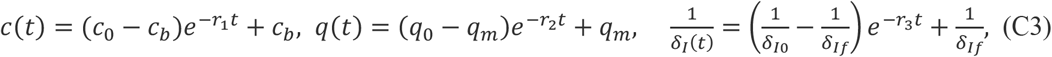

where *c*_0_ denotes the initial contact rate (on January 23^rd^ 2020) with *c*(0) = *c*_0_, *c*_*b*_ denotes the minimal contact rate with the intervention being implemented. Constant *q*_0_ is the initial quarantined rate of exposed individuals with *q*(0) = *q*_0_, *q*_*m*_ is the maximum quarantined rate with *q*_*m*_ > *q*_0_. Similarly, constant δ_*I*0_ is the initial diagnose rate, δ_*If*_ is the fastest diagnose rate with δ_*If*_ > δ_*I*0_.

For the Guandong province, the recovery rate has been improved over the time, and this is reflected by the time-dependence of the recovery rate of the quarantined infected (isolated) population, given by

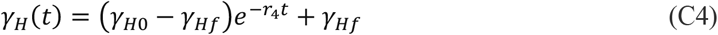

with *γ*_*H*0_ being the initial recovery rate and *y*_*Hf*_ the fastest recovery rate, and *R*_1_ the corresponding increasing rate of the recovery rate.

For South Korea, a similar contact tracing strategy has been implemented: all traced individuals are quarantined and are tested. Once tested, the suspected individuals leave the compartment at a rate *b*, with a proportion, *f*, if confirmed to be COVID-19 infected, going to the hospitalized compartment, whilst the other proportion, 1-*f*, is recommended to remain quarantined and hence goes to the compartment *S*_*q*_^18,19^. The transmission dynamics subject to this intervention practice is given by

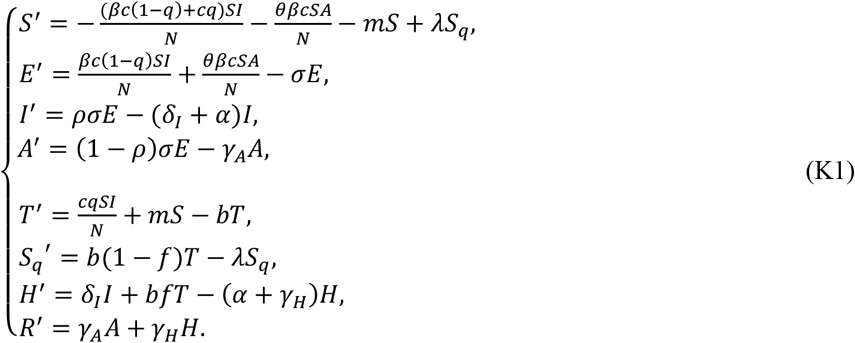

The basic reproduction number is given by

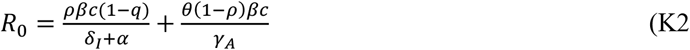

### Model-free estimation for *R*_0_ and *R*_*t*_

We employed the method developed by White and Pagano^20^ to estimate the basic reproduction number *R*_0_. 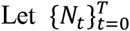 be the number of newly reported cases on day 0,…,T. Assume N_*t*_ follows the Poisson distribution with a mean of 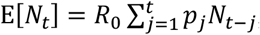, where *p*_j_(j = 1,2, …) is the probability, giving the serial interval distribution. Assume also that N_*I*_ and N_j_ are independent as long as *I* ≠ j. Given the distribution of the serial interval and the observed data on 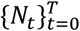, the basic reproduction number *R*_0_ can be estimated by the maximum likelihood estimates approach.

With the same notations and assumptions described above, we can also estimate the effective reproduction number *R*_*t*_ following Cori^21^. Namely, using 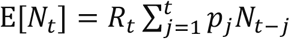 within the Bayesian framework, we can obtain an analytical expression of the posterior distribution of *R*_*t*_ by assuming a gamma prior distribution for *R*_*t*_. Then we can get the posterior means and confidence intervals of *R*_*t*_.

## Results

### Model-free estimation for *R*_0_ and *R*_*t*_

By using the number of daily newly reported cases from January 10^th^ to January 23^rd^ 2020, we estimate *R*_0_ for mainland China, and using the newly reported cases from January 19^th^ to January 31^st^ 2020 we estimate *R*_0_ for the Guangdong province. Also, we estimate *R*_0_ for South Korea based on the number of daily newly reported cases from January 23^rd^ to March 2^nd^ 2020.

All the estimates are given in Table 2. In particular, given the serial interval with mean of 5 and standard deviation of 3, *R*_0_ for mainland China, the Guangdong Province and South Korea is estimated to be 3.8 (95%CI: (3.5, 4.2)), 3.0 (95%CI: (2.6, 3.3)) and 2.6 (95%CI: (2.5, 2.7)), respectively. In particular, the initial COVID-19 reproduction rate in South Korea was smaller than that in the Guangdong Province.

**Table 1:** Parameter estimates for the COVID-19 epidemic in China, Guangdong Province of China and Korea.

| Parameter |  | Definitions | Estimated values |  |  | Source |
| --- | --- | --- | --- | --- | --- | --- |
|  |  |  | Guangdong | China | Korea |  |
| $c(t)$ | $c_0$ | Contact rate at the initial time | 10.00(Est) | 14.78 | -- | Data |
| | $c_b$ | Minimum contact rate under the current control strategies | 4.00 | 7.78 | -- | Estimated |
| | $r_1$ | Exponential decreasing rate of contact rate | 0.01 | 2 | -- | Estimated |
| $c$ | | Constant contact rate | -- | -- | 20.4121 | Estimated |
| $\beta$ | | Probability of transmission per contact | 0.13 | 0.143 | 0.143(F) | Estimated |
| $\theta$ | | Modification factor of the transmission probability of asymptotical infected individual | 0.02 | 0.0437 | 0.1539 | Estimated |
| $q(t)$ | $q_0$ | Quarantined rate of exposed individuals at the initial time | 0.28 | $1.00 \times 10^{-4}$ | -- | Estimated |
| | $q_m$ | Maximum quarantined rate of exposed individuals under the current control strategies | 0.99 | 0.8 | -- | Estimated |
| | $r_2$ | Exponential increasing rate of quarantined rate of exposed individuals | 0.0361 | 0.15 | -- | Estimated |
| $q$ | | Constant quarantined rate | -- | -- | 0.7480 | Estimated |
| $m$ | | Transition rate of susceptible individuals to the suspected class | $1 \times 10^{-5}$ | $1 \times 10^{-6}$ | $1.1 \times 10^{-4}$ | Estimated |
| $b$ | | Detection rate of the suspected class | 0.0891 | 0.20 | 0.2144 | Estimated |
| $f$ | | Confirmation ratio: Transition rate of exposed individuals in the suspected class to the quarantined infected class | 0.01 | 0.50 | 0.0624 | Estimated |
| $\rho$ | | Ratio of symptomatic infection | 0.5 | 0.4022 | 0.8409 | Estimated |
| $\sigma$ | | Transition rate of exposed individuals to the infected class | 1/5 | 1/5 | 1/5 | Data |
| $\lambda$ | | Rate at which the quarantined uninfected contacts were released into the wider community | 1/14 | 1/14 | 1/14 | Data |
| $\delta_I(t)$ | $\delta_{I0}$ | Initial transition rate of symptomatic infected individuals to the quarantined infected class | 0.12(Est) | 0.1326 | -- | Data |
| | $\delta_{If}$ | Fastest diagnose rate | 0.50 | 0.50 | -- | Estimated |
| | $r_3$ | Exponential decreasing rate of diagnose rate | 0.2410 | 0.10 | -- | Estimated |
| $\delta_I$ | | Constant transition rate of symptomatic infected individuals to the quarantined infected class | -- | -- | 0.1131 | Estimated |
| $\gamma_A$ | | Recovery rate of asymptotic infected individuals | 0.1397 | 0.1397 | 0.1397 | Data |
| $\gamma_H(t)$ | $\gamma_{H0}$ | Recovery rate of quarantined infected individuals at initial time | 0.001 | -- | -- | Estimated |
| | $\gamma_{Hf}$ | Fastest recovery rate of quarantined infected individual | 0.2283 | -- | -- | Estimated |
| | $r_4$ | Exponential increasing rate of recovery rate of quarantined infected individuals | 0.01 | -- | -- | Estimated |

| $\gamma_H$ | Constant recovery rate of quarantined infected individuals | -- | 0.2 | 0.2(F) | Estimated |
| --- | --- | --- | --- | --- | --- |
| $\alpha$ | Disease-induced death rate | 0(Assumed) | 0.0076 | 0.076(F) | Estimated |
| Initial values | Definitions | Estimated values |  |  | Source |
|  |  | Guangdong | China | Korea |  |
| $S(0)$ | Initial susceptible population | $8.00 \times 10^5$ | $3.00 \times 10^7$ | $2.01 \times 10^6$ | Estimated |
| $E(0)$ | Initial exposed population | 250 | $3.00 \times 10^3$ | 177.44 | Estimated |
| $I(0)$ | Initial infected population | 378 | $4.00 \times 10^3$ | 19.811 | Estimated |
| $A(0)$ | Initial asymptotical population | 100 | $1.00 \times 10^3$ | 34.106 | Estimated |
| $T(0)$ | Initial suspected population | 258 | 1072 | $1.03 \times 10^3$(Est) | Data |
| $S_q(0)$ | Initial quarantined susceptible population | 21(Est) | 7347 | 7123 | Data |
| $H(0)$ | Initial quarantined infected population | 53 | 771 | 29 | Data |
| $R(0)$ | Initial recovered population | 2 | 34 | 0 | Data |

Note that, ‘Est’ means that the parameter values are estimated by fitting the models to the data when the source column indicates that they are not. ‘F’ means that the parameter values are fixed as the same as those estimated by fitting the proposed model to the multiple source data of China.

**Table 2:** Estimated basic reproduction number *R*_0_ for mainland China, Guangdong province of China, Korea for various serial intervals.

| $R_0(95\% \text{ CI})$ | | | | | |
| --- | --- | --- | --- | --- | --- |
| **Std=3** | **E=4** | **E=5** | **E=6** | **E=7** | **E=8** |
| **Mainland China** | 2.9(2.6,3.2) | 3.8(3.5,4.2) | 5.4(4.9,6.0) | 7.8(7.1,8.6) | 11.1(10.0,12.2) |
| **Guangdong** | 2.4(2.1,2.7) | 3.0(2.6,3.3) | 3.8(3.4,4.3) | 5.1(4.5,5.8) | 6.9(6.1,7.8) |
| **South Korea** | 2.1(2.0,2.2) | 2.6(2.5,2.7) | 3.4(3.2,3.5) | 4.5(4.3,4.7) | 6.0(5.7,6.3) |
| **Std=4** | **E=4** | **E=5** | **E=6** | **E=7** | **E=8** |
| **Mainland China** | 2.8(2.5,3.1) | 3.4(3.1,3.7) | 4.4(4.0,4.9) | 6.0(5.4,6.6) | 8.4(7.6,9.2) |
| **Guangdong** | 2.4(2.1,2.7) | 2.8(2.5,3.1) | 3.4(3.0,3.9) | 4.4(3.9,4.9) | 5.8(5.1,6.5) |
| **South Korea** | 2.1(2.0,2.1) | 2.4(2.3,2.5) | 3.0(2.8,3.1) | 3.8(3.7,4.0) | 5.0(4.8,5.3) |
| **Std=5** | **E=4** | **E=5** | **E=6** | **E=7** | **E=8** |
| **Mainland China** | 2.8(2.5,3.1) | 3.2(2.9,3.5) | 3.9(3.5,4.3) | 4.9(4.5,5.6) | 6.6(5.9,7.2) |
| **Guangdong** | 2.4(2.1,2.7) | 2.7(2.4,3.0) | 3.2(2.8,3.6) | 3.9(3.4,4.3) | 4.9(4.3,5.5) |
| **South Korea** | 2.1(2.0,2.2) | 2.3(2.2,2.4) | 2.7(2.6,2.9) | 3.4(3.2,3.5) | 4.3(4.1,4.4) |

E: mean of serial interval, Std: standard deviation of serial interval.

In order to investigate the variation of *R*_0_ with respect to the serial interval, we carry out a sensitivity analysis by changing the mean of the serial interval from 4 to 8 days, the standard deviation (Std) from 3 to 5. The sensitivity analysis is reported in Table 2, and we notice that *R*_0_ increases when the mean of the serial interval increases and we remark that serial interval examined by recent studies is shorter than that earlier estimation^22-24^. It also follows from Table 2 that increasing Std of the serial interval only slightly decreases the estimated *R*_0_ for a given mean of the serial interval.

We also estimate the effective reproduction numbers *R*_*t*_ for the considered regions, using the number of daily newly reported cases from the date the first case was reported until March 2^nd^ 2020 (Figure 3). It shows that the effective reproduction number *R*_*t*_ in mainland China and in its Guandong province has fallen below the threshold 1 since February 18^th^ and February 7^th^, while the effective reproduction number of South Korea remains very high, indicating that there is still room for improving the interventions in South Korea.

**Figure 3:**
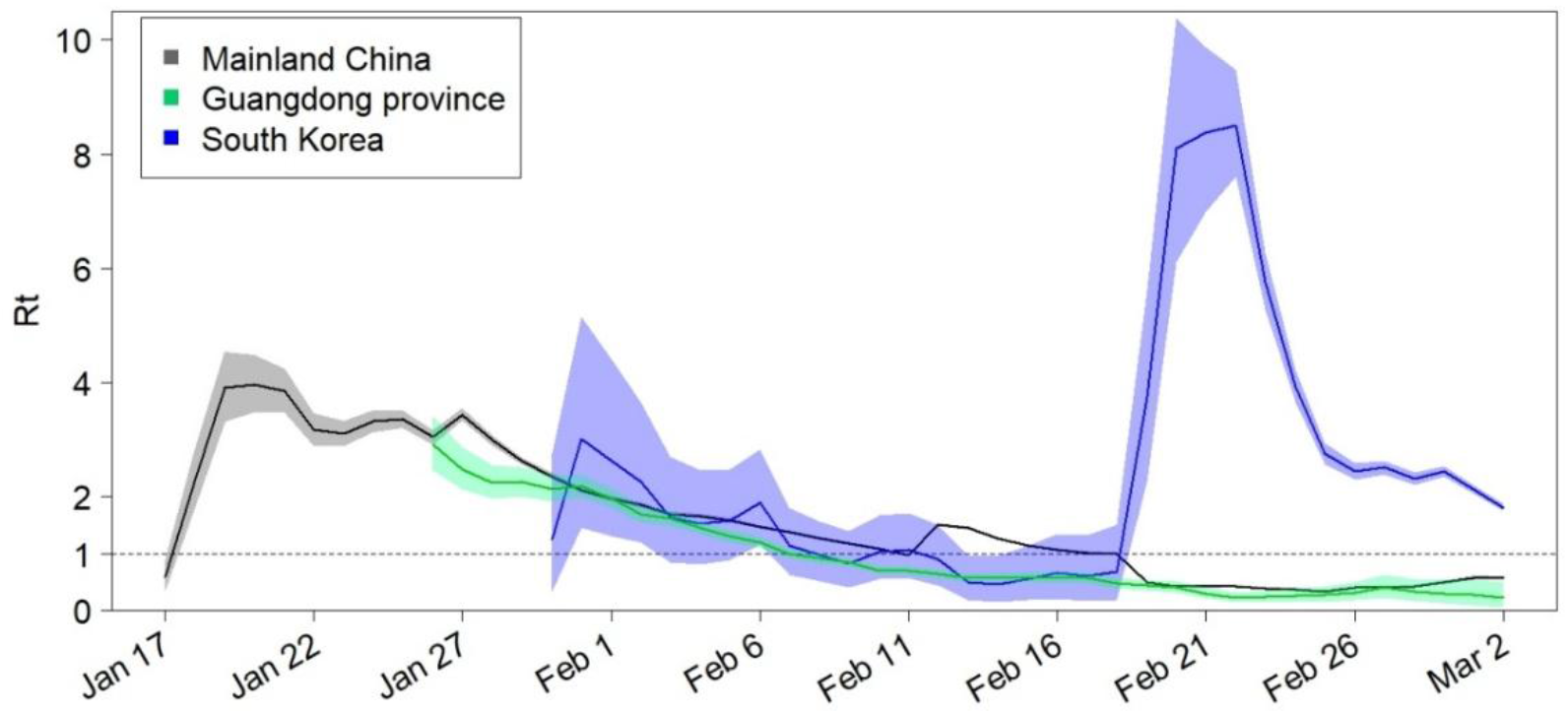
Estimated effective reproduction number *R*_*t*_ over sliding weekly windows for the entire country of China, the Guangdong province of China, and Korea. The solid lines show the posterior means and the coloured zones show the 95% confidence intervals; the horizontal dashed line indicates *R*_*t*_=1.

### Model-based prediction and interventions efficacy evaluation

By simultaneously fitting the model (C1) to the multiple source data on the cumulative number of reported cases, deaths, quarantined and suspected cases in mainland China, we obtain estimations for unknown parameters and initial conditions, listed in Table 1. The best fitting result is shown as black curves in Figure 5 with the estimated baseline exponential decreasing rate (*r*_1_ = *r*_10_) in the contact rate function. We then conduct a sensitivity analysis of the cumulative reported, death, quarantined, suspected cases, and the infected (asymptomatic/symptomatic) individuals by shrinking the exponential index *r*_1_, representing the weakening of the control interventions relevant to the contact rate. As shown in Figure 5, the numbers of cumulative reported, death, quarantined, suspected cases, and the peak value of the infected all increase significantly. In particular, with *r*_1_ = 0 corresponding to no reduction of the contact rate from the initial period, the cumulative confirmed cases increases by more than six times as of April 1^st^ (∼600,000 cases) and the peak value of the infected will increase by more than 3 times, in comparison with the actual situation under the strong control measures implemented by the Chinese government.

**Figure 5.**
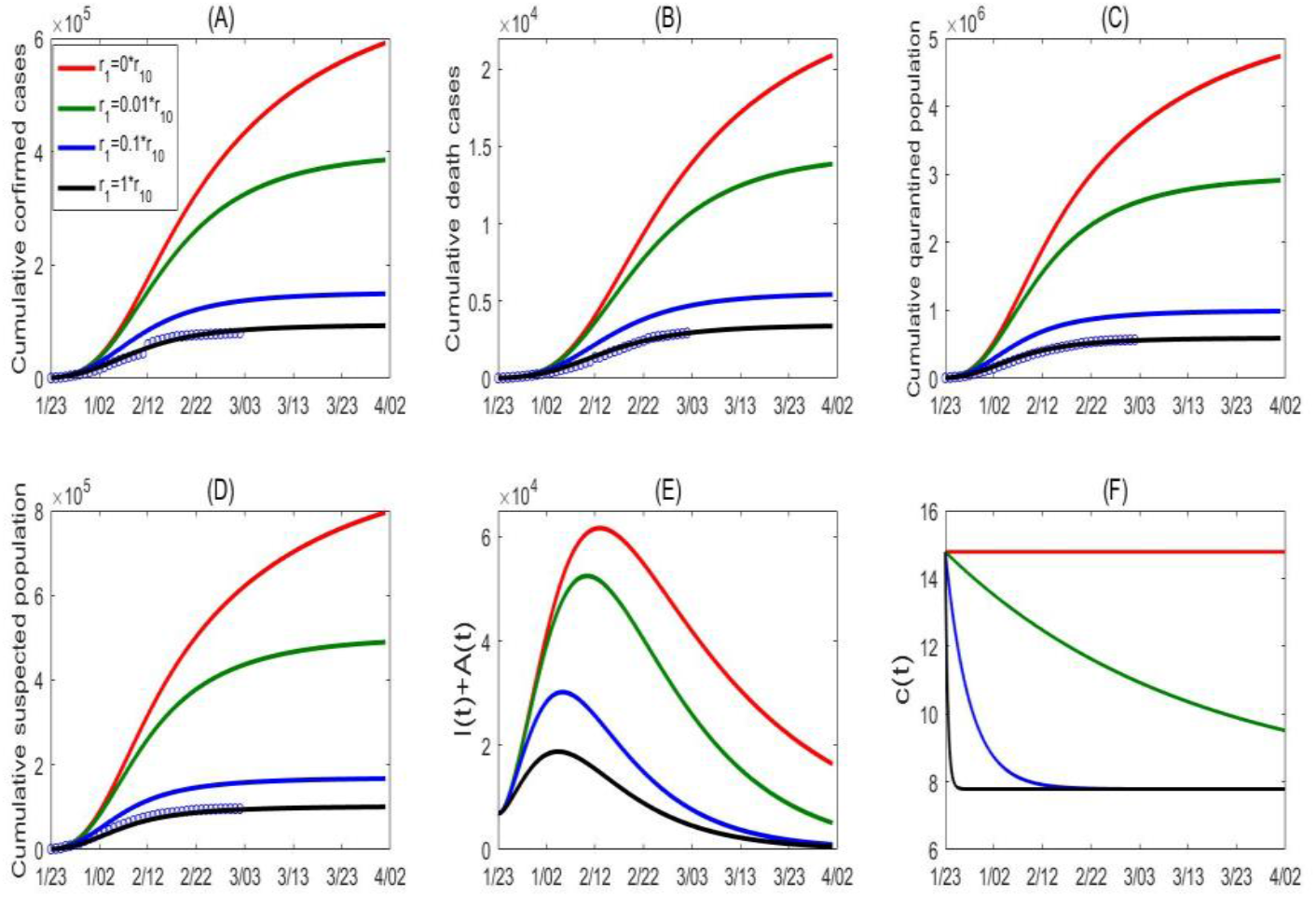
Goodness of fit (black curves) and variations in cumulative number of reported cases (A), death cases (B), quarantined cases (C) and suspected cases (D) for mainland China. (E) Variation in number of infected (asymptomatic/symptomatic) individuals with contact rate function *c*(*t*). (F) Here the contact rate function is changed by varying the exponential decreasing rate *r*_1_, representing the variation in intensity of control measures. *r*_10_ denotes the estimated baseline value of *r*_1_.

We also conduct a sensitivity analysis regarding the detection rate *δ*_*I*_ (*t*), by decreasing the value of *r*_3_. We obtain a similar conclusion that the cumulative confirmed cases would reach the number of 350,000 cases as of April 1^st^ with a constant detection rate (no improvement of detection), shown in Figure 6. As illustrated in Figure 6(F), we can observe that while decreasing the detection rate would not affect the decreasing trend of the effective reproduction number, it however postpones the time when the threshold value of 1 is reached. Therefore the outcome in the mainland China, both in terms of the infections avoided and the timing when the outbreak begins to be under control, is the consequence of a systematic package of social distancing (self-isolation and self-protection), contacting tracing, and detection/diagnosis.

**Figure 6.**
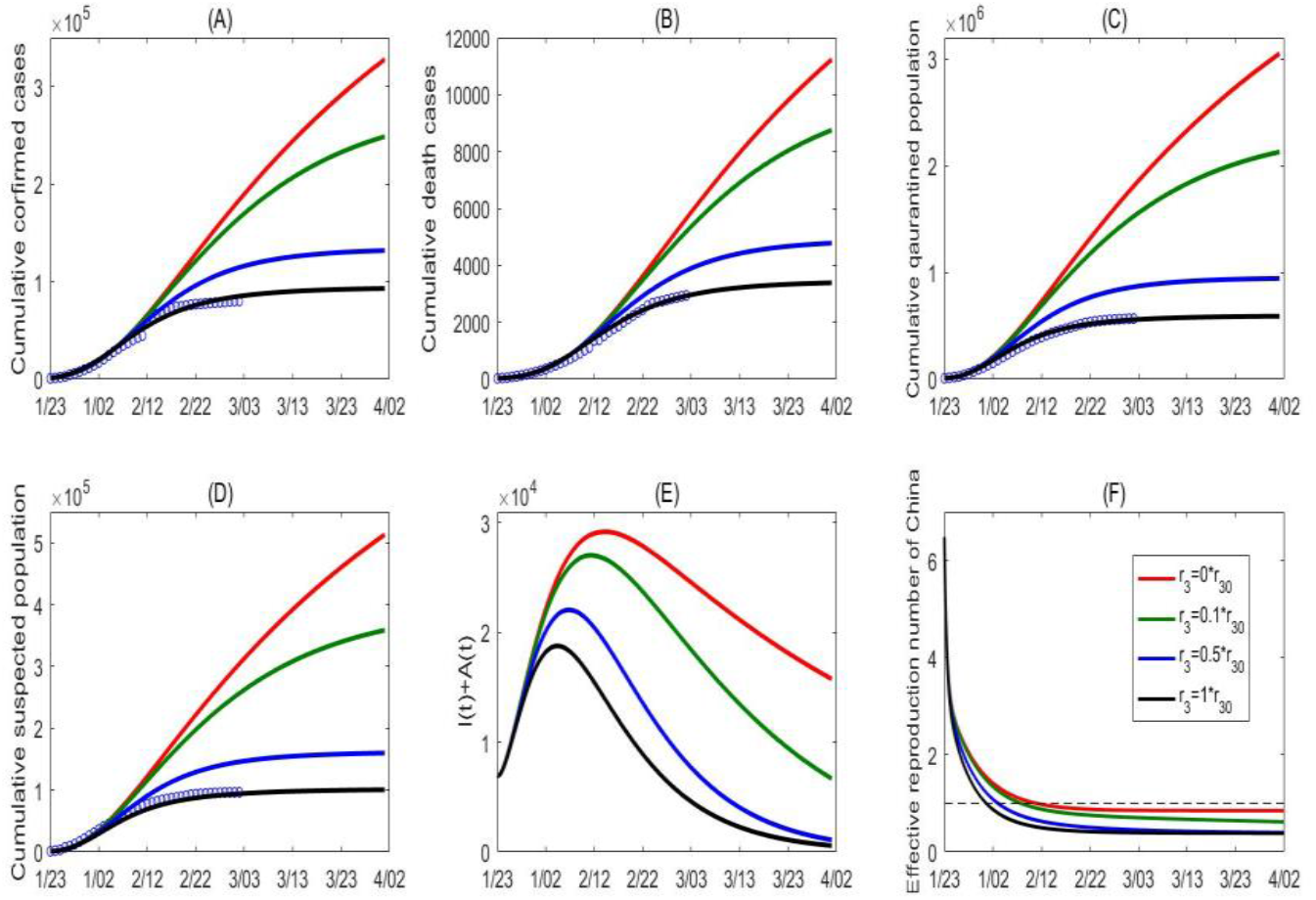
Goodness of fit (black curves) and variations in cumulative number of reported cases (A), cumulative number of death cases (B), quarantined cases (C) and suspected cases (D) for mainland China. (E) Variation in number of infected (asymptomatic/symptomatic) individuals with detection rate function *δ*_*I*_ (*t*). (F) Variation in the effective reproduction number of China. Here the detection rate function is changed by varying the exponential decreasing rate *r*_3_, representing the variation in intensity of control measures. *r*_30_ denotes the estimated baseline value of *r*_3_.

Similarly, by simultaneously fitting the proposed model (K1) to the cumulative number of reported and tested cases for South Korea, we obtain the estimations for the unknown parameters and initial conditions, listed in Table 1. The best fitting result is shown as black curves in Figure 7 with the estimated constant contact, testing and detection rates. For the purpose of a comparative study, we simulate the situation in South Korea by importing some of the interventions and measures implemented in the mainland China. We focus on the cases when we can 1) replace the contact rate and detection rate estimated in the (K1) model from the South Korea data with the time-dependent rate function (C3), and 2). adopt the time-dependent testing rate similarly to the quarantined rate function q(t) in (C3) to use

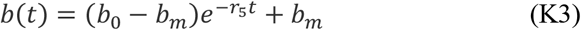

with parameters *b*_0_, *b*_*m*_ to indicate the initial and maximal testing rate. We report the simulations in four scenarios:

Scenario A: Using the testing rate function in (K3), representing an enhanced testing strategy, the cumulative tested and confirmed cases significantly increase, and the cumulative confirmed cases will reach 300 thousands on April 5^th^ 2020 (red curves, Figure 7(C-D)).
Scenario B: Using only the detection rate function in (C3), the cumulative tested and confirmed cases increase too, and the cumulative confirmed cases will reach 200 thousands on April 5^th^ 2020 (green curves, Figure 7(C-D)).
Scenario C: Using only the contact rate function in (C3), the cumulative confirmed cases will reach 100 thousands on April 5^th^ 2020 (blue curves, Figure 7(C-D)).
Scenario D: Using the time-dependent contact, detection and testing rate functions, representing an integrated systematic package of public health control strategies, the cumulative confirmed cases will reach around 60 thousands on April 5^th^ 2020 (black curves, Figure 7(C-D)).

**Figure 7.**
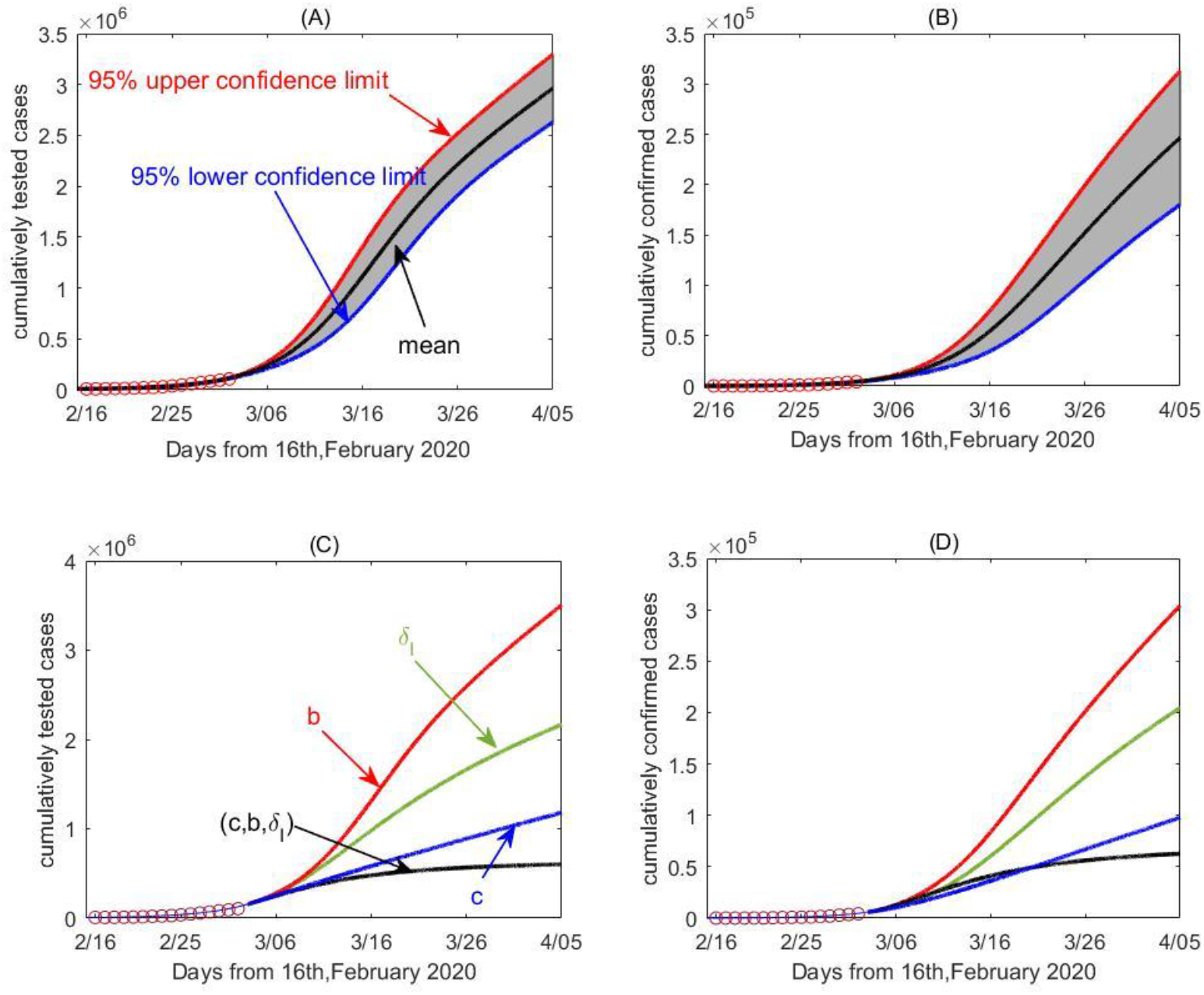
The impact of the randomness of the cumulative reporting data sets including cumulative number of reported cases, cumulative number of death cases, cumulative quarantined cases and cumulative suspected cases on the 2019nCov epidemic in mainland China. The unilateral 95% confidence intervals (here 95% upper confidence limits) have been given, and the mean curve and estimated curve based on the real data sets are marked in each subplot.

We conclude that a significant reduction of COVID-19 cases is achievable only through a systematic package consisting of enhanced control measures including self-isolation/self-production, effective quarantine and rapid detection/testing.

Similarly, by simultaneously fitting the model (C1) to the multiple source data on the cumulative number of reported cases, recovery and suspected cases of Guangdong province, we parameterize the model and obtain the estimations for the unknown parameters and initial conditions, listed in Table 1. The best fitting result is shown as green curves in Figure 8. Similarly, we consider the variation in the epidemic by means of the parameter ***r***_**1**_, representing the variation in the intensity of the control measures implemented. It follows from Figure 8(A-C) that increasing the parameter ***r***_**1**_ reduces the disease infections and the number of suspected individuals, while decreasing the parameter ***r***_**1**_ (to zero) slightly affects the disease infections, shown in green curves (***r***_**1**_ ≠ **0**) and red curves (***r***_**1**_ = **0**) in Figure 8(A-C). This conversely implies that the prevention and control strategies in Guangdong province were relatively strong from the early stage of the outbreak. This can also be confirmed by the continuously declining trend of the effective reproduction number, as shown in Figure 3 and 8(D). Again, we performed the sensitivity analysis by decreasing the value of *r*_**3**_ in the detection rate ***δ***_***I***_(***t***) and obtained similar results, as shown in Figure 9.

**Figure 8.**
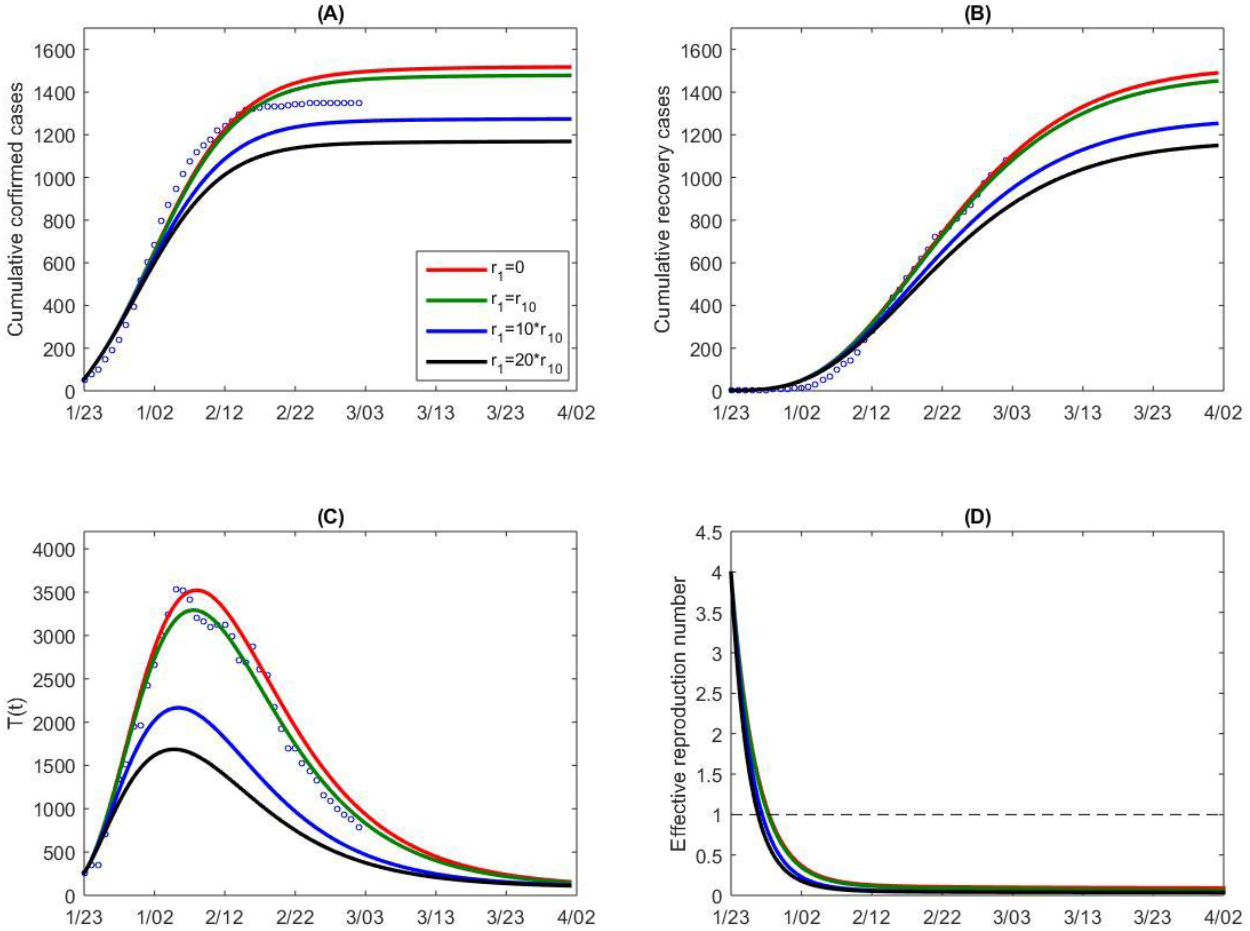
Goodness of fit (green curves) and variations in cumulative number of reported cases (A), recovery cases (B), and suspected cases (C) for Guangdong province. (D) Variation in the effective reproduction number with parameter *r*_1_ in contact rate function *c*(*t*). Here the contact rate function is changed by varying the exponential decreasing rate *r*_1_, representing the variation in intensity of control measures. *r*_10_ denotes the estimated baseline value for parameter *r*_1_.

**Figure 9.**
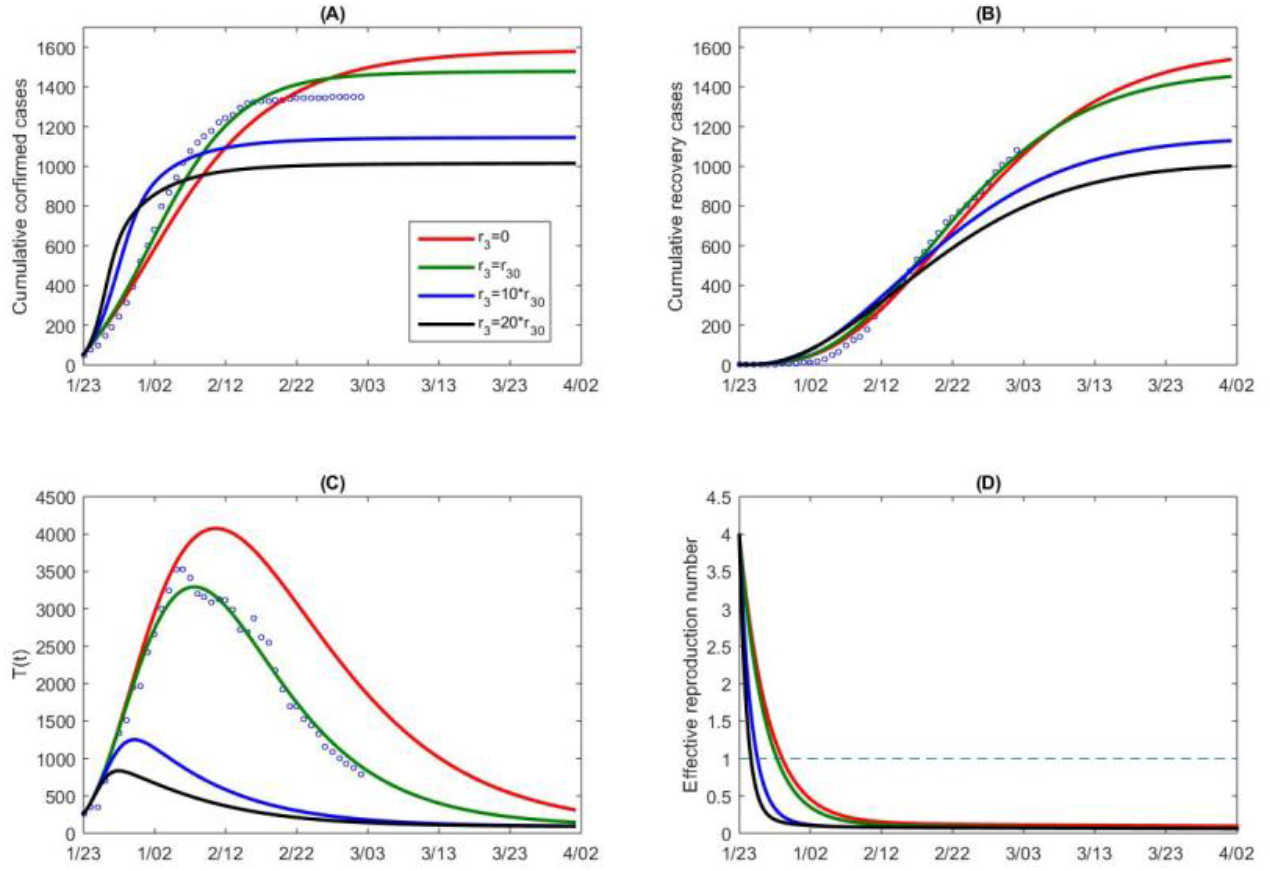
Goodness of fit (green curves) and variations in cumulative number of reported cases (A), recovery cases (B), and suspected cases (C) for Guangdong province. (D) Variation in the effective reproduction number with parameter *r*_3_ in detection rate function *δ*_*I*_ (*t*). Here the detection rate function is changed by varying the exponential decreasing rate *r*_3_, representing the variation in intensity of control measures. *r*_30_ denotes the estimated baseline value of *r*_3_.

## Discussion

After some initial delay in the response to the COVID-19 outbreak, the Chinese government started implementing drastic public health measures on January 23^rd^ 2020, including the lock-down of Wuhan city, contact tracing/quarantine/isolation. This decisive and systematically implemented package of interventions has contributed to rapid mitigation, providing a window of opportunity for the preparedness, prevention and control of COVID-19 in other Chinese cities and around the world. As we showed in this modeling study, a systematic approach incorporating coherent and complementary public health measures rapidly implemented is the key for containing the viral outbreak.

In this study, we formulated a dynamic model and parameterized it by using multi-source data for different settings, like South Korea, China and Guandong province. We predicted that the trend of cumulative reported cases would be very serious, reaching around 600 thousands as of April 1^st^ 2020 given no control measures being implemented. Hence a comparison with the current COVID-19 epidemic: the cumulative reported cases are around 80,651 as of March 6^th^, 2020, indicating the efficacy of the ongoing strengthening of prevention and control measures in mainland China.

In Guangdong, the province with the largest population in China, most cases at the initial phase of the COVID-19 outbreak are imported, and immediately after the lockdown of Wuhan on January 23^rd^ 2020, the province implemented a systematic approach towards prevention and control with gradual enhancement leading to the effective control of otherwise potentially catastrophic outcomes.

In comparison with South Korea, Guangdong has more inhabitants and a less developed economy. Also, from our model-free estimation, the basic reproduction number in South Korea is less than that computed for the Guangdong province. Therefore, the COVID-19 epidemic potential in South Korea was initially weaker than that in Guangdong. However, our model-based analysis also shows that the effective reproduction number in South Korea remains greater than 1 while the epidemic in the Guangdong province has already been under control. Our simulation results indicate that the COVID-19 epidemic in South Korea will change from a quick to a slow increase if the integrated control measures are implemented, as illustrated in Figure 7(C). Hence, the experience of epidemic control in mainland China is worth popularizing, especially for the reference of Western countries and other settings, including South Korea.

More in detail, a comparison of the parameter estimations of the Guangdong province and the entire country China shows that

1. the initial and maximum quarantine rates in Guangdong were much higher than those in the entire country China, while the initial and minimum contact rates were lower than those in the country, contributing to the observed better control effect in the province than the national average.
2. the confirmation ratios of the Guangdong province and South Korea were much lower than the ratio of the entire country China, indicating the better efficiency of contact tracing and testing in the Guangdong province and South Korea than that in the entire country of China.
3. the constant contact rate in South Korea was larger than that in the Guangdong province and even the entire country of China, with an even bigger minimum contact rate in South Korea, suggesting the need of raising the awareness of the importance of self-isolation and self-protection.

Carrying out the sensitivity analysis of confirmed cases of China and Guangdong province with respect to the contact rate function, this suggests the effectiveness of the control measures implemented in South Korea. Other countries or territories experiencing large-scale outbreaks of COVID-19 should capitalize on this lesson: convincing all the citizens to stay at home by self-isolation and become aware of the implications of self-protection is of crucial importance. Similarly, the detection rate *δ*_*I*_ of Korea is also lower than those of China and Guangdong province. Therefore, improving the diagnose rate of the infectious individual is also fundamental for controlling the spreading of COVID-19 epidemic in Korea.

## Data Availability

Data utilized are publicly available

## Author Contributions

Conceptualization, S.T., Y.X. and J.W.; methodology, S.T., Y.X. and J.W.; software, B.T., F.X., X.W.X.S. and S.T.; validation, S.T., Y.X. and J.W.; formal analysis, B.T., X.F.,X.W. X.S. and S.T.; investigation, S.T., Y.X. and J.W.; resources, B.T., X.F., X.W. and S.T.; data curation, B.T., X.F., X.W. and S.T.; writing—original draft preparation, B.T., X.F., X.W., N.L.B. and S.T.; writing—review and editing, N.L.B., S.T., Y.X. and J.W.; visualization, J.W.; supervision, S.T. and J.W.; project administration, J.W.; funding acquisition, Y.X. and J.W. All authors have read and agreed to the published version of the manuscript.

